# Tumor Suppressor Immune Gene Therapy to Reverse Immunotherapy Resistance

**DOI:** 10.1101/2020.12.27.20248913

**Authors:** Sunil Chada, Dora Wiederhold, Kerstin B. Menander, Beatha Sellman, Max Talbott, John J. Nemunaitis, Hyo Min Ahn, Bo-Kyeong Jung, Chae-Ok Yun, Robert E. Sobol

## Abstract

**Background:** While immune checkpoint inhibitors are becoming a standard of care for multiple types of cancer, the majority of patients do not respond to this form of immunotherapy. New approaches are required overcome resistance to immunotherapies.

**Methods:** We investigated the effects of adenoviral p53 (Ad-p53) gene therapy in combination with immune checkpoint inhibitors and selective IL2 or IL15 CD122/132 agonists in the aggressive B16F10 tumor model resistant to immunotherapies. To assess potential mechanisms action, pre and post Ad-p53 treatment biopsies were evaluated for changes in gene expression profiles by Nanostring IO 360 assays.

**Results:** Substantial synergy of “triplet” Ad-p53 + CD122/132 + anti-PD-1 therapy resulted in potential curative effects associated with complete tumor remissions of both primary and contralateral tumors. Interestingly, contralateral tumors which were not injected with Ad-p53 showed robust abscopal effects resulting in statistically significant decreases in tumor size and increased survival (p<0.001). None of the monotherapies or doublet treatments induced complete tumor regressions. Ad-p53 treatment increased Type I Interferon, CD8^+^ T cell, immuno-proteosome antigen presentation and tumor inflammation gene signatures. Ad-p53 treatment also decreased immune suppressive TGF-beta, beta-catenin, macrophage, and endothelium gene signatures, which may contribute to enhanced immune checkpoint inhibitor (CPI) efficacy. Unexpectedly, a number of previously unidentified, strongly p53 down regulated genes associated with stromal pathways and IL10 expression identified novel anti-cancer therapeutic applications.

**Conclusions:** These results imply the ability of Ad-p53 to induce efficacious local and systemic anti-tumor immune responses with the potential to reverse resistance to immune checkpoint inhibitor therapy when combined with CD122/132 agonists and immune checkpoint blockade. Our findings further imply that Ad-p53 has multiple complimentary immune mechanisms of action which support future clinical evaluation of triplet Ad-p53, CD122/132 agonist and immune checkpoint inhibitor combination treatment.

## Background

Immune checkpoint inhibitor therapy has become a new standard of care for multiple recurrent and metastatic cancers. However, most cancer patients do not respond to this form of treatment(1). Various approaches are being tested to increase immune checkpoint blockade efficacy including combination with immune stimulating cytokine(2, 3). Interleukin 2 (IL2) and interleukin 15 (IL15) belong to a family of immune stimulating cytokines sharing a common beta chain (CD122) and gamma-chain (CD132) receptor known to drive the proliferation and cytolytic activity of CD8+ T cells and natural killer (NK) cells. Selective CD122/132 agonists have been developed with minimal alpha chain (CD25) binding that mitigate generation of capillary leak toxicities and the induction of immune suppressive T regs(4-7).

TP53 is the prototypic tumor suppressor that regulates responses to a wide range of cell stressors including cell cycle arrest, cellular senescence, apoptosis, DNA damage repair, hypoxia, oncogenic stress and epithelial-mesenchymal transition (EMT)(8). Ad-p53 is a replication impaired adenoviral vector encoding expression of the wild-type p53 tumor suppressor protein which has demonstrated anti-tumor effects in pre-clinical and clinical studies as a monotherapy and in combination with other treatment modalities (9-13).

We evaluated Ad-p53 tumor suppressor therapy in a murine tumor model known to be highly resistant to immunotherapy in combination with IL2/IL15 CD122/132 agonists and immune checkpoint blockade. We observed substantial synergy supporting further development of triplet Ad-p53, CD122/132 agonist and immune checkpoint inhibitor combination treatment.

## Methods

### Animals, tumor inoculation and measurements

C57BL/6 (B6) mice (6-8 weeks of age) were injected subcutaneously into the right flank with B16F10 melanoma cells (ATCC, 5 x 10^5^ cells/mouse) to form the “Primary Tumor”. Treatment started when tumors reached approximately 60 mm^3^ in size (designated Day 1). On day 14, animals were inoculated on the contralateral side with B16F10 cells to form “Secondary tumor”, and primary and secondary tumor growth followed for up to 60 days. Animals were sacrificed when tumors reached approximately 2000 mm^3^.

### Viral vectors

Replication-deficient human type 5 adenovirus (Ad5) encoding for expression of p53 tumor suppressor gene was used. The construction, properties and purification of the vector have been reported elsewhere for Ad-p53 vectors(11). Four doses of viral vectors were administered, intra-tumorally, at 48-hour intervals (5 x 10^9^ viral particles/dose, in 50 µl total volume).

### Anti-PD-1 Treatment

Animals were treated with intraperitoneal anti-PD-1(10mg/kg) every 3 days starting on Day 1, over a 30-day period following implantation of the primary tumor. Outcome measures were subcutaneous tumor growth, measured twice per week by caliper measurements. Tumor growth was monitored by measuring the length (L) and width (w) of the tumor, and tumor volume calculated using the following formula: volume = 0.523L(w)^2^. Treatment outcomes were evaluated by measurement of tumor volumes in primary and contralateral tumors and their statistical analyses by T test, analysis of variance (ANOVA), Kruskal-Wallis ANOVA; and by comparisons of survival using Kaplan-Meier and log rank tests.

### CD122/ CD132 Agonist Treatment

Murine IL-2 (eBioscience or R&D Systems Minneapolis, MN) was mixed with the S4B6-1 anti-mouse IL2 antibody (Bioxcell, West Lebanon, NH or BD Biosciences) at a molar ratio 2:1 to generate the preferential CD122/CD132 agonist immunocomplex. For studies involving human T cells, human IL-2 was mixed with MAB602 anti-human IL-2 antibody (R&D Systems). The IL-2/S4B6 or IL-2/MAB602 mAb immunocomplexes were administered intraperitoneally (IP) at 2.5 µg IL2/dose on days 2, 6, and 10. Alternatively, IL-2/S4B6 mAb immunocomplexes were injected on days 2– 6 (1.0 µg IL2/dose). Immunocomplexes are prepared by incubating anti-IL-2 monoclonal with IL-2 for 15 minutes at room temperature.

In some experiments, the CD122/CD132 agonist was comprised of recombinant mouse IL-15 (eBiosciences) and IL-15-R alpha-Fc (R&D Systems). The immunocomplex was prepared by incubated these together at 37 C for 30 minutes, and this preferential CD122/CD132 agonist immunocomplex was injected i.v. for two consecutive days once tumors become palpable. An alternate schedule is administration of the IL-15 immune complex injected IP on days 3, 5 and 7 after tumors become palpable. For IL-15 immunocomplex studies, recombinant murine IL-15 (Peprotech, Rocky Hill, CT, USA) is used in the in vivo studies at doses of 2 µg/injection recombinant murine IL-15 once per week by intravenous injection. Recombinant mouse IL-15 R alpha Fc Chimera Protein is obtained from R&D Systems (Minneapolis, MN) and used at doses equimolar to IL-15 cytokine (12 µg/injection of IL-15-Ra-Fc for each 2 ug IL-15 protein in immune complex).

### Nanostring Transcriptome Gene Expression Analyses of Ad-p53 Treatment

We also describe in this report, the initial transcriptome results of gene expression profiles induced by Ad-p53 treatment performed as part of a new Ad-p53 recurrent HNSCC clinical trial in combination anti-PD-1 therapy NCT03544723 (https://clinicaltrials.gov/ct2/show/NCT03544723). RNA was isolated from pre- and post-treatment samples and compared using Nanostring IO 360 gene expression panel (Nanostring Technologies Seattle WA). This panel tests expression of 770 genes involved in neoplasm pathology, tumor microenvironment and cancer immune responses. Samples were processed and analyzed as described(14). mRNA expression was measured with the nCounter technology, provided by NanoString Technologies. nCounter uses probes with barcodes attached to DNA oligonucleotides that directly bind to RNA. Sample preparation and analyses were performed according to the manufacturer’s protocol using The PanCancer IO 360 gene expression panel that includes 770 genes. Gene expression signatures were defined as described previously(14, 15). Normalization was performed by correcting for the expression of technical controls and 30 housekeeping genes included in the panel. nCounter gene expression data were obtained for pre-and post-treatment biopsies.

### Statistical Analysis

Graph Pad Prism 8.0 software was employed for statistical analyses. A statistical analysis of variance (ANOVA) was employed to compare treatment effects on tumor size. For survival comparisons, Kaplan-Meier survival estimates, and the log rank test were utilized. Fisher’s exact test was employed for comparison of complete remission rates. Statistically significant p values were less than or equal to 0.05.

## Results

### Ad-p53 Has Local and Abscopal Effects with Reversal of Anti-PD-1 Resistance

We evaluated the ability of Ad-p53 to reverse resistance to immune checkpoint inhibitor therapy in the B16F10 melanoma tumor model, which is known to be refractory to immunotherapy. Mimicking clinical applications, we allowed tumors to progress on anti-PD-1 treatment before intra-tumoral administration of Ad-p53 therapy to tumors in one flank with contralateral tumors not injected with Ad-p53. Local treatment effects on the Ad-p53 injected tumor are shown in Figure 1. Ad-p53 + anti-PD-1 combination treatment induced statistically significant decreases in tumor growth compared to either anti-PD-1 or Ad-p53 therapy alone. Evaluation of tumor growth using ANOVA statistical analysis confirmed synergistic effects of the combination treatment over either agent used as monotherapy (p=0.0001). We observed only minimal anti-PD-1 monotherapy efficacy with results similar to the tumor progression seen for control PBS treatment. In contrast, treatment with Ad-p53 monotherapy resulted in significantly decreased tumor growth and Ad-p53 + anti-PD-1 combination therapy reversed anti-PD-1 treatment resistance (See Figure 1). Treatment with a combination of Ad-luciferase (Ad-Luc) and anti-PD-1 did not enhance the effect of anti-PD-1 therapy (see Supplemental Figure 1).

**Figure 1.**
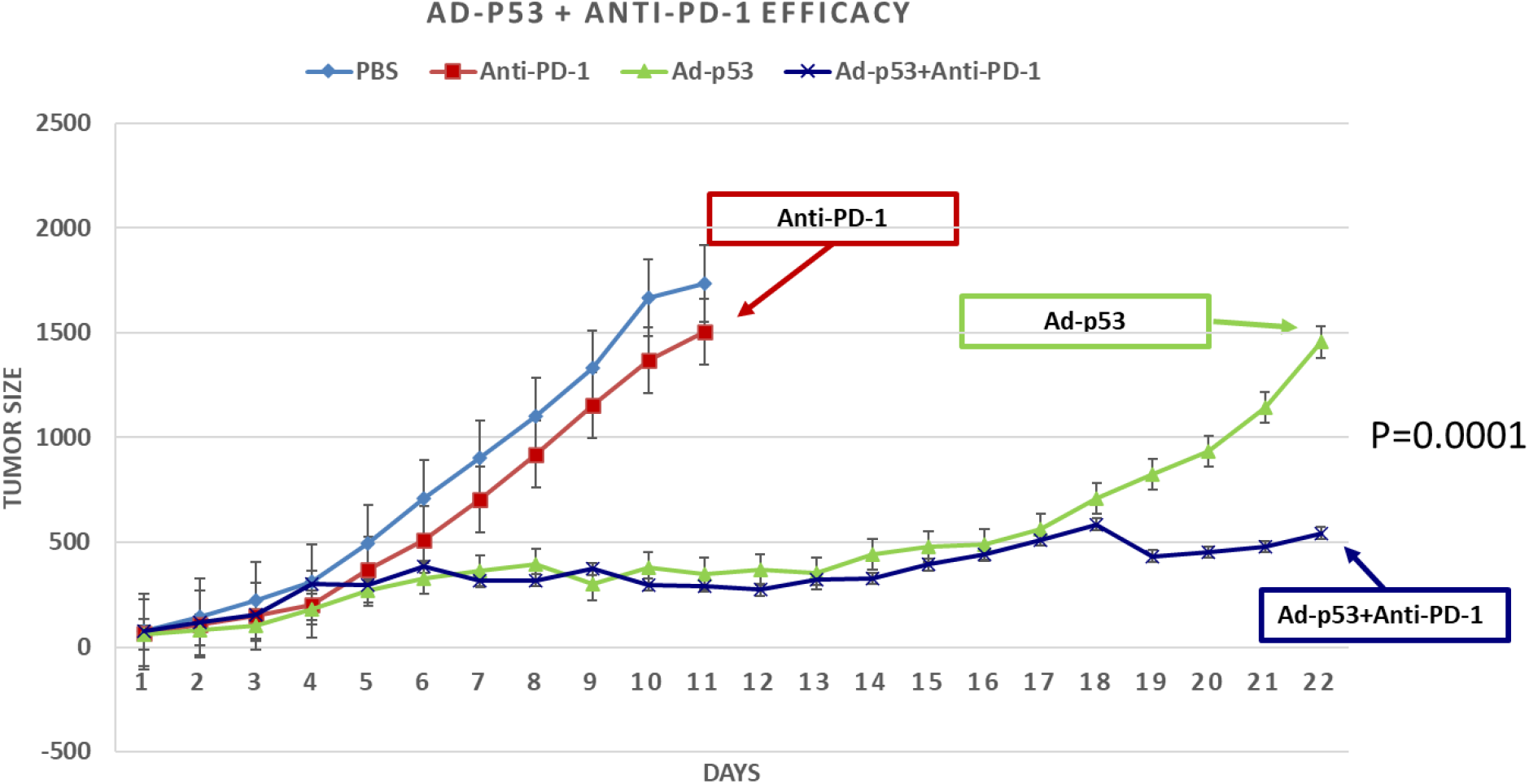
Local Effects on Ad-p53 Injected Tumors + anti-PD-1 Therapy. Ad-p53 + anti-PD-1 combination treatment induced statistically significant decreases in tumor growth compared to either anti-PD-1 or Ad-p53 therapy alone. Evaluation of tumor growth using ANOVA statistical analysis confirmed synergistic effects of the combination treatment over either agent used as monotherapy (p=0.0001). Anti-PD-1 monotherapy had minimal efficacy with substantial tumor progression similar to animals control PBS treatment. Treatment with Ad-p53 monotherapy resulted in significantly delayed tumor growth and Ad-p53 + anti-PD-1 combination therapy reversed anti-PD-1 treatment resistance.

As shown in Figure 2, abscopal, systemic anti-tumor effects of localized Ad-p53 treatment were observed in contralateral tumors that were not injected with Ad-p53. Consistent with the synergistic effect seen in the suppression of Ad-p53 injected tumors, we also observed a statistically significant abscopal effect with decreased growth in the contralateral tumors that did not receive Ad-p53 tumor suppressor therapy. These findings imply that the combination treatment (Ad-p53 + anti-PD1) induced systemic immunity mediating the abscopal effects. Contralateral tumors in animals whose primary tumor had been treated with Ad-p53 alone showed significantly delayed tumor growth (p=0.046) compared to the growth rate of primary tumors treated with anti-PD-1 alone. An even greater abscopal effect on contralateral tumor growth (p=0.0243) was observed in mice whose primary tumors were treated with combined Ad-p53+anti-PD-1.

**Figure 2.**
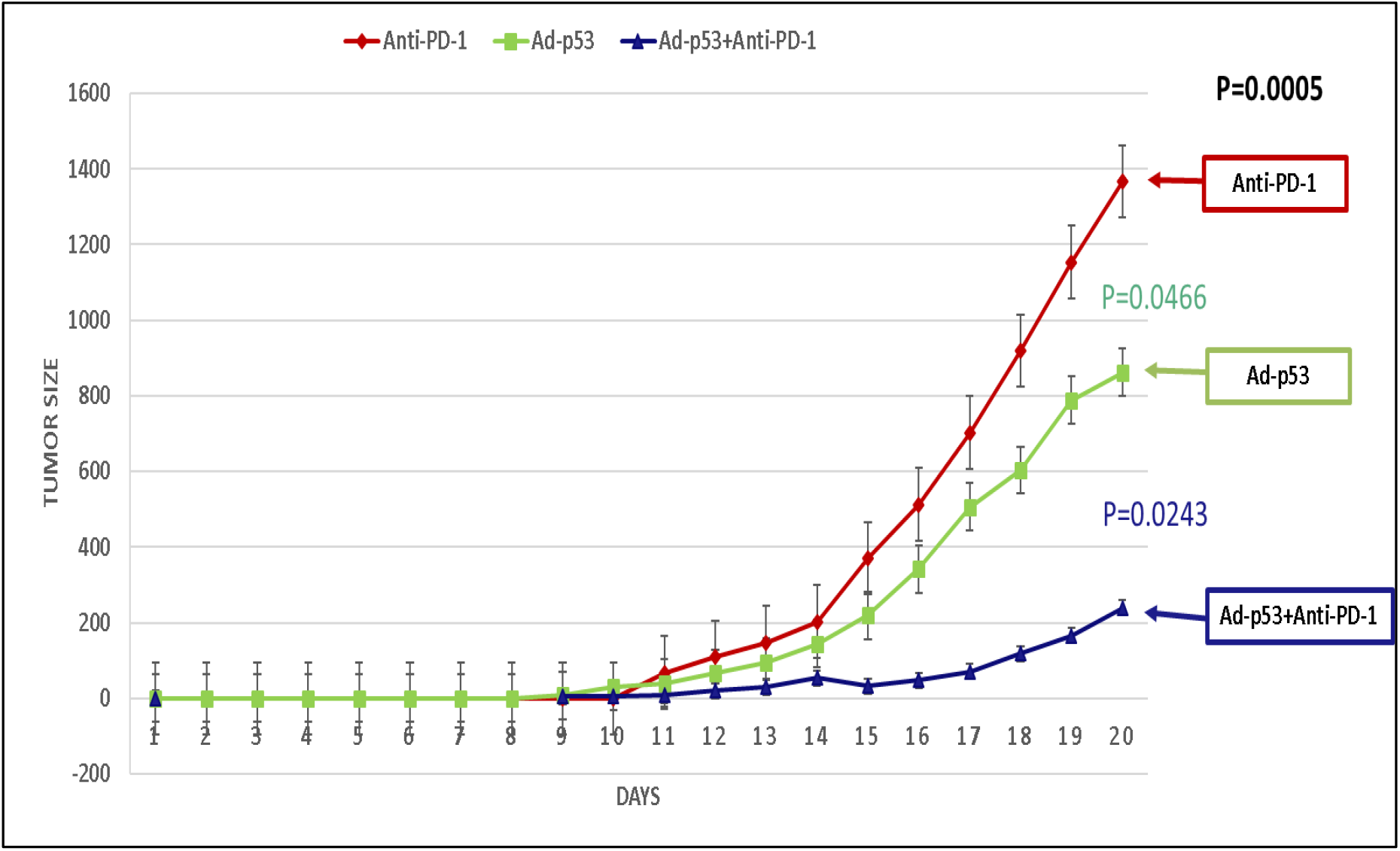
Abscopal Effect of Ad-p53 in contralateral tumors not injected with Ad-p53. Contralateral tumor volume over time in rodents whose primary tumor had received either anti-PD-1, Ad-p53 or a combination of Ad-p53 + anti-PD-1 treatment. Consistent with the synergistic effect observed in the suppression of primary tumor growth, we also observed a statistically significant abscopal effect with decreased growth in the contralateral (secondary) tumors that did not receive tumor suppressor therapy. These findings imply that the combination treatment (Ad-p53 + anti-PD1) induced systemic immunity mediating the abscopal effects. Contralateral tumors in animals whose primary tumor had been treated with Ad-p53 alone showed significantly delayed tumor growth (p=0.046) compared to the growth rate of primary tumors treated with anti-PD-1 alone. An even greater abscopal effect on contralateral tumor growth (p=0.0243) was observed in mice whose primary tumors were treated with combined Ad-p53+anti-PD-1.

With respect to survival, combined Ad-p53 and anti-PD-1 therapy demonstrated a statistically significant increase in survival compared to Ad-p53 therapy alone (p = 0.0167) and anti-PD-1 therapy alone (p < 0.001) (see Figure 3). Consistent with the synergistic effects on tumor growth, the increase in median survival for the combined Ad-p53 and anti-PD-1 group was more than additive compared to the effects of Ad-p53 and anti-PD-1 treatments.

**Figure 3.**
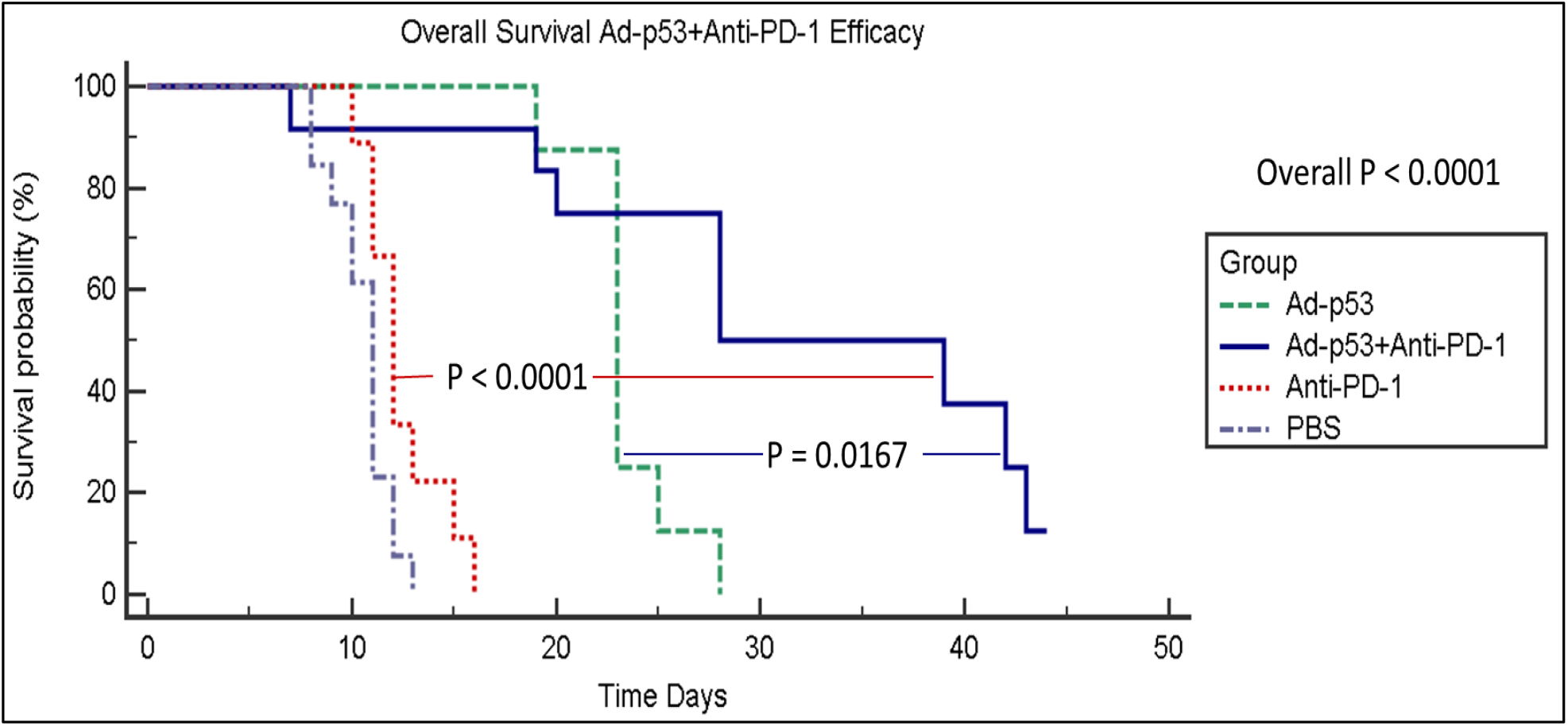
Kaplan-Meier survival curves showing superior efficacy for Ad-p53 + anti-PD-1 therapy. Kaplan-Meier survival curves for mice treated with either PBS, anti-PD-1, Ad-p53 or a combination of these agents. The results show no significant difference in the survival of animals treated with PBS or anti-PD-1, increased survival in those treated with Ad-p53, and a significant enhancement of survival in animals treated with a combination of Ad-p53 + anti-PD-1 over that observed in mice treated with either Ad-p53 (p=0.0167), or anti-PD-1 (p<0.001) monotherapy.

### Local and Systemic Efficacy by “Triplet” Ad-p53 + Immune Checkpoint Blockade + CD122/133 Agonist Therapy

In a subsequent series of experiments, we evaluated the effects of combining IL2 or IL15 CD122/CD132 agonists with Ad-p53 tumor suppressor and immune checkpoint blockade. Treatment efficacy was evaluated by assessing tumor volumes (in primary and contralateral tumors), complete tumor response rates and survival. The results demonstrated unexpected, substantial synergy of the “Triplet”Ad-p53 + CD122/132 + anti-PD-1 therapies that resulted in potentially curative treatment associated with complete tumor remissions of both primary and contralateral tumors with significantly superior abscopal effects on distant tumors not injected with Ad-p53 tumor suppressor therapy. As demonstrated in Figures 4, 5 and 6, the triplet Ad-p53 + CD122/132 + anti-PD-1 therapy was the only treatment that resulted in complete tumor remissions and long term survival.

**Figure 4.**
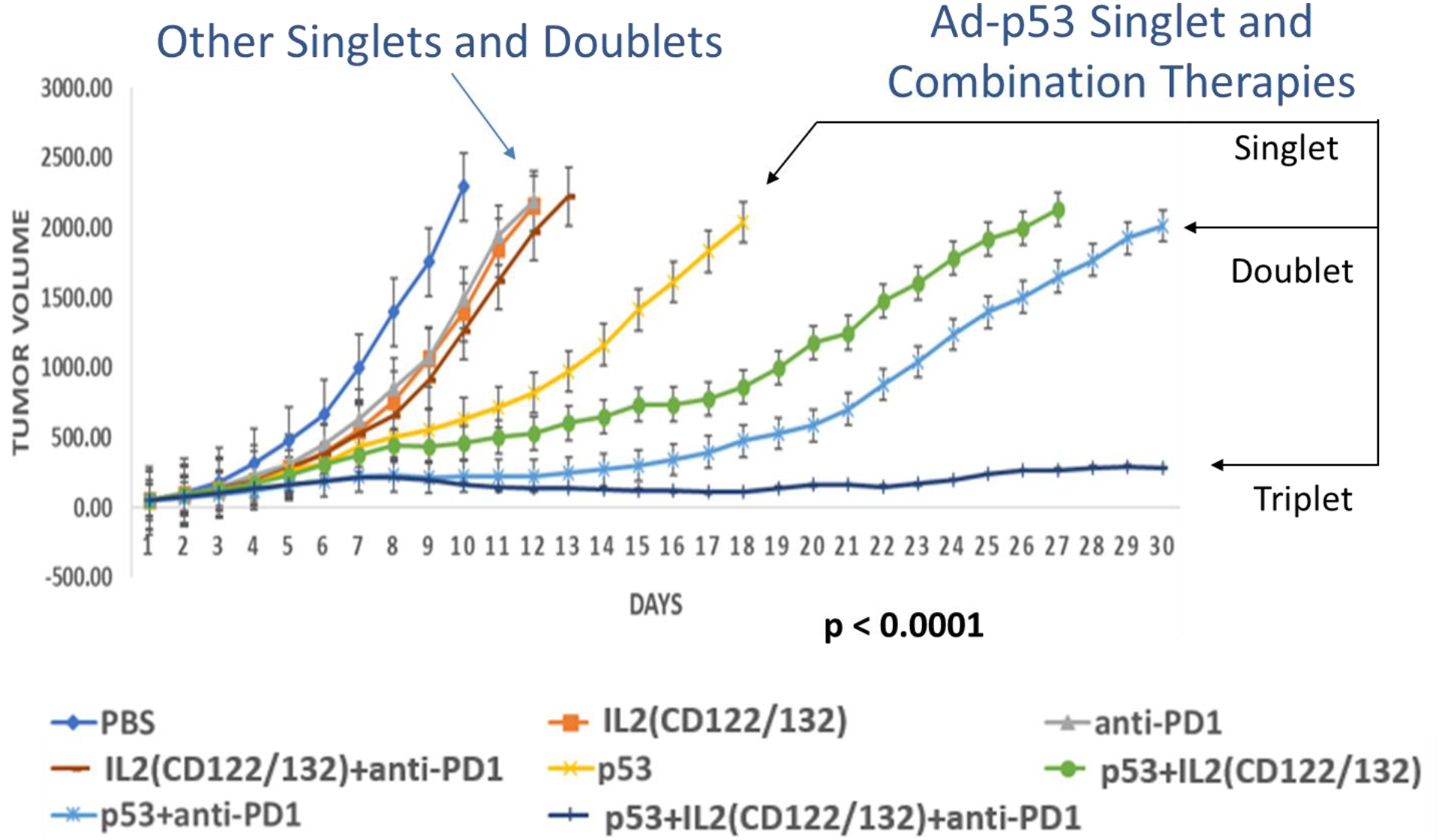
Substantially Superior Efficacy of “Triplet” Ad-p53 + IL2 CD122/132 agonist + anti-PD-1 Therapy. There was significantly enhanced efficacy of Triplet Ad-p53 + CD122/132 + anti-PD-1 treatment compared to any of the other singlet or doublet therapies. Importantly, a statistical analysis of variance (ANOVA) comparison of tumor volumes on Day 30 determined that the synergy of the anti-tumor effects was only maintained in the “Triplet” Ad-p53 + CD122/132 + anti-PD-1 treatment combination (p-value < 0.0001 overall and p-value < 0.0001 separately compared to every other treatment group).

**Figure 5.**
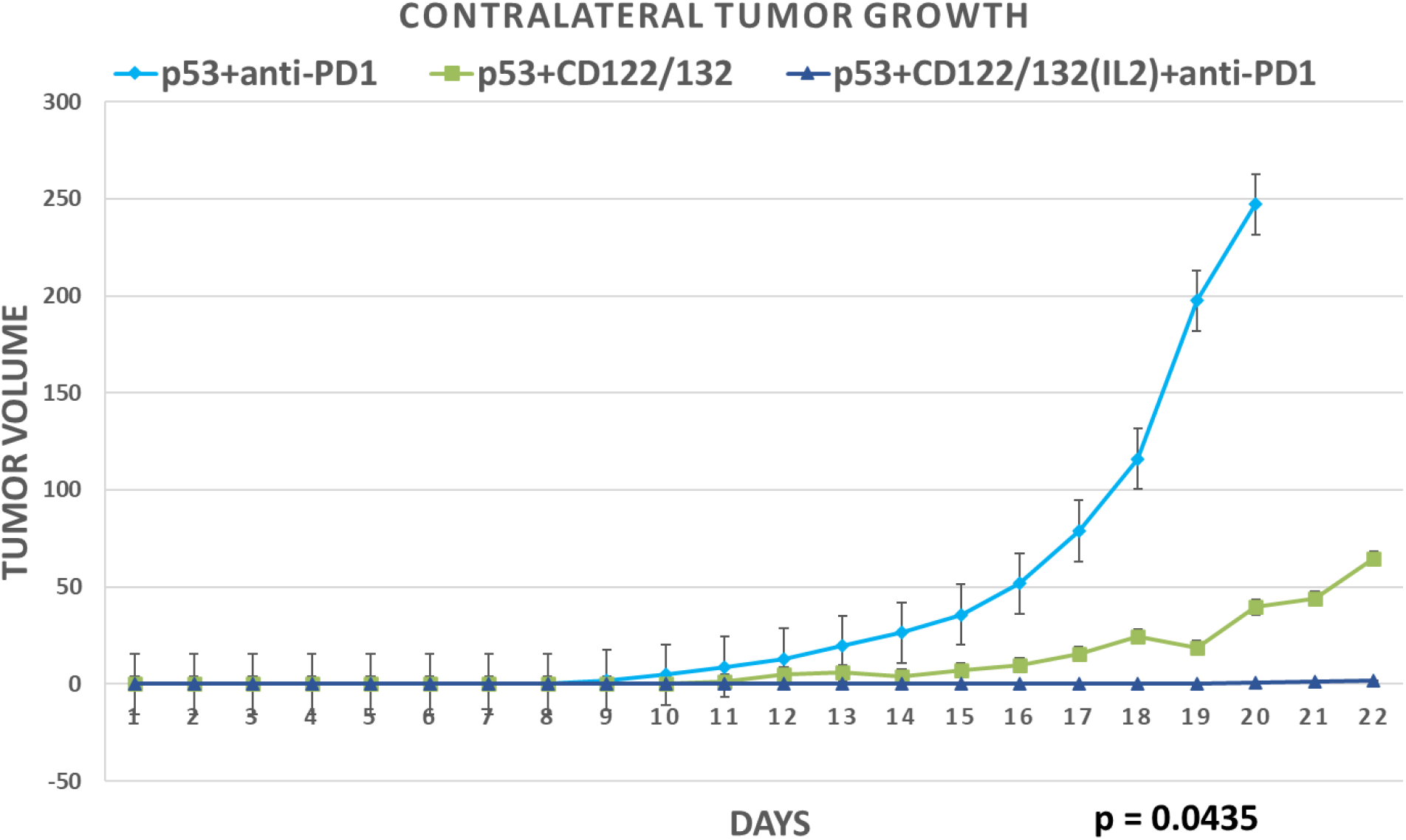
Substantially Superior Abscopal/Systemic Efficacy of “Triplet” Ad-p53 + IL2 CD122/132 agonist + anti-PD-1 Therapy. Consistent with the unexpected, substantially increased synergistic effects of Ad-p53 + CD122/132 + anti-PD-1 treatment on primary tumor growth, we also observed a surprisingly powerful and statistically significant abscopal effect of triplet Ad-p53 + CD122/132 + anti-PD-1 treatment compared to the other Ad-p53 treatment groups. A statistical analysis of variance (ANOVA) comparison of these contralateral tumor volumes determined synergy of the anti-tumor effects of Ad-p53 + CD122/132 + anti-PD-1 treatment (p-value = 0.0435 overall). Only the Ad-p53 + CD122/132 + anti-PD-1 group demonstrated a statistically significant decrease in contralateral tumor growth vs. the Ad-p53 + anti-PD-1 group (p-value = 0.0360).

**Figure 6.**
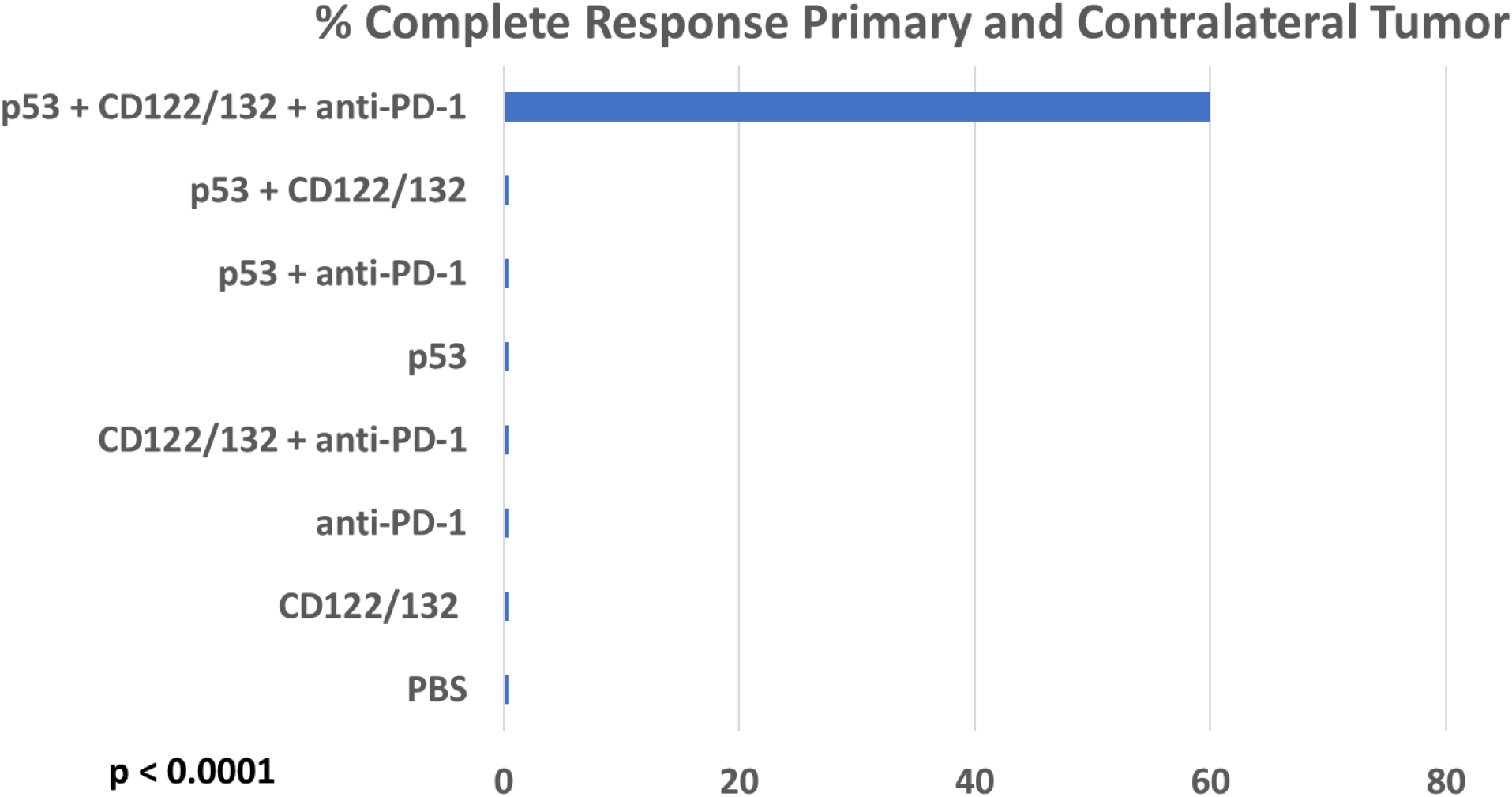
“Triplet” Ad-p53 + CD122/132 + anti-PD-1 treatment induces complete tumor responses. Only Ad-p53 + CD122/132 + anti-PD-1 treatment resulted in complete tumor remissions of both primary and contralateral tumors. Complete tumor responses of both primary and contralateral tumors were observed in 60% of the Ad-p53 + CD122/132 + anti-PD-1 treatment group (6 of 10 animals) and there were no complete tumor responses in any of the 70 animals in the other treatment groups (p-value < 0.0001 by two-sided Fisher’s Exact Test comparing Ad-p53 + CD122/132 + anti-PD-1 treatment group vs. animals in all other treatment groups; p-value < 0.011 by two-sided Fisher’s Exact Test comparing Ad-p53 + CD122/132 + anti-PD-1 treatment group vs. any other treatment group).

### Significant Improvement of Local Efficacy of “Triplet” Ad-p53 + Immune Checkpoint Blockade + CD122/133 Agonist Therapy in Primary Tumor Injected with Ad-p53

In regard to primary tumor volume as shown in Figure 4, there was enhanced efficacy of Ad-p53 + CD122/132, Ad-p53 + anti-PD-1 and Ad-p53 + CD122/132 + anti-PD-1 treatments compared to any of the therapies alone. Importantly, a statistical analysis of variance (ANOVA) comparison of tumor volumes on Day 30 determined that the synergy of the anti-tumor effects was only maintained in the “Triplet” Ad-p53 + CD122/132 + anti-PD-1 treatment combination (p-value < 0.0001 overall and p-value < 0.0001 separately compared to every other treatment group). There was severe tumor progression during CD122/132, anti-PD-1, and CD122/132 + anti-PD-1 therapies which were reversed by combination with Ad-p53 therapy.

### Superior Abscopal/Systemic Effects of “Triplet” Ad-p53 Tumor Suppressor Immune Therapy

As shown in Figure 5, abscopal, systemic anti-tumor effects in contralateral tumors that were not injected with Ad-p53 were significantly superior for Ad-p53 “Triplet” therapy. Figure 5 depicts contralateral tumor volumes over time in rodents receiving the three most effective tumor treatments with either the combination of Ad-p53 + IL2 CD122/132, Ad-p53 + anti-PD-1, or Ad-p53 + IL2 CD122/132 + anti-PD-1. A statistical analysis of variance (ANOVA) comparison of these contralateral tumor volumes determined synergy of the anti-tumor effects of Ad-p53 + IL2 CD122/132 + anti-PD-1 treatment (p-value = 0.0435). Similar results were observed with IL15 derived CD122/132 treatments (See Supplemental Figure 2).

### “Triplet” Ad-p53 + CD122/132 + anti-PD-1 treatment induces complete tumor responses

It is generally appreciated that complete tumor responses to therapy are associated with important therapeutic benefits and are required for curative outcomes. As shown in Figure 6 for the p53 treatment groups and their controls, only Ad-p53 + CD122/132 + anti-PD-1 treatment resulted in complete tumor remissions of both primary and contralateral tumors. Complete tumor responses of both primary and contralateral tumors were observed in 60% of the Ad-p53 + CD122/132 + anti-PD-1 treatment group (6 of 10 animals) and there were no complete tumor responses in any of the 70 animals in the other treatment groups (p-value < 0.0001 by two-sided Fisher’s Exact Test comparing Ad-p53 + CD122/132 + anti-PD-1 treatment group vs. animals in all other treatment groups; p-value < 0.011 by two-sided Fisher’s Exact Test comparing Ad-p53 + CD122/132 + anti-PD-1 treatment group vs. any other treatment group). Unexpectedly, the complete tumor responses were durable and were maintained after 40 days in 50% of the Ad-p53 + CD122/132 + anti-PD-1 treatment group presumably curing these animals of these tumors. Taken together, these findings indicate that of all the Ad-p53 therapies, only the triplet combination Ad-p53 + CD122/132 + anti-PD-1 treatment resulted in curative efficacy by inducing powerful local and systemic anti-tumor immunity mediating substantial abscopal effects.

### “Triplet” Ad-p53 + CD122/132 + anti-PD-1 treatment results in extended survival

The Kaplan-Meier survival curves shown in Figure 7 demonstrate the substantial synergy of triplet Ad-p53 + CD122/132 + anti-PD-1 therapy compared to mice treated with either PBS or Ad-p53 monotherapy, or the doublet therapies CD122/132 + anti-PD-1, Ad-p53 + CD122/132 or Ad-p53 + anti-PD-1. There was a statistically significant difference in these survival curves by the log rank test (p<0.0001 overall; p-value < 0.0003 comparing Ad-p53 + CD122/132 + anti-PD-1 treatment group vs. any other treatment group). The median survival of the triplet Ad-p53 + CD122/132 + anti-PD-1 therapy group had not been reached after 40 days and 80% of this treatment group were still alive. In stark contrast, 98% (49/50) of animals in the other treatment groups had died by Day 30 and had median survivals ranging between 10 to 28 days.

**Figure 7.**
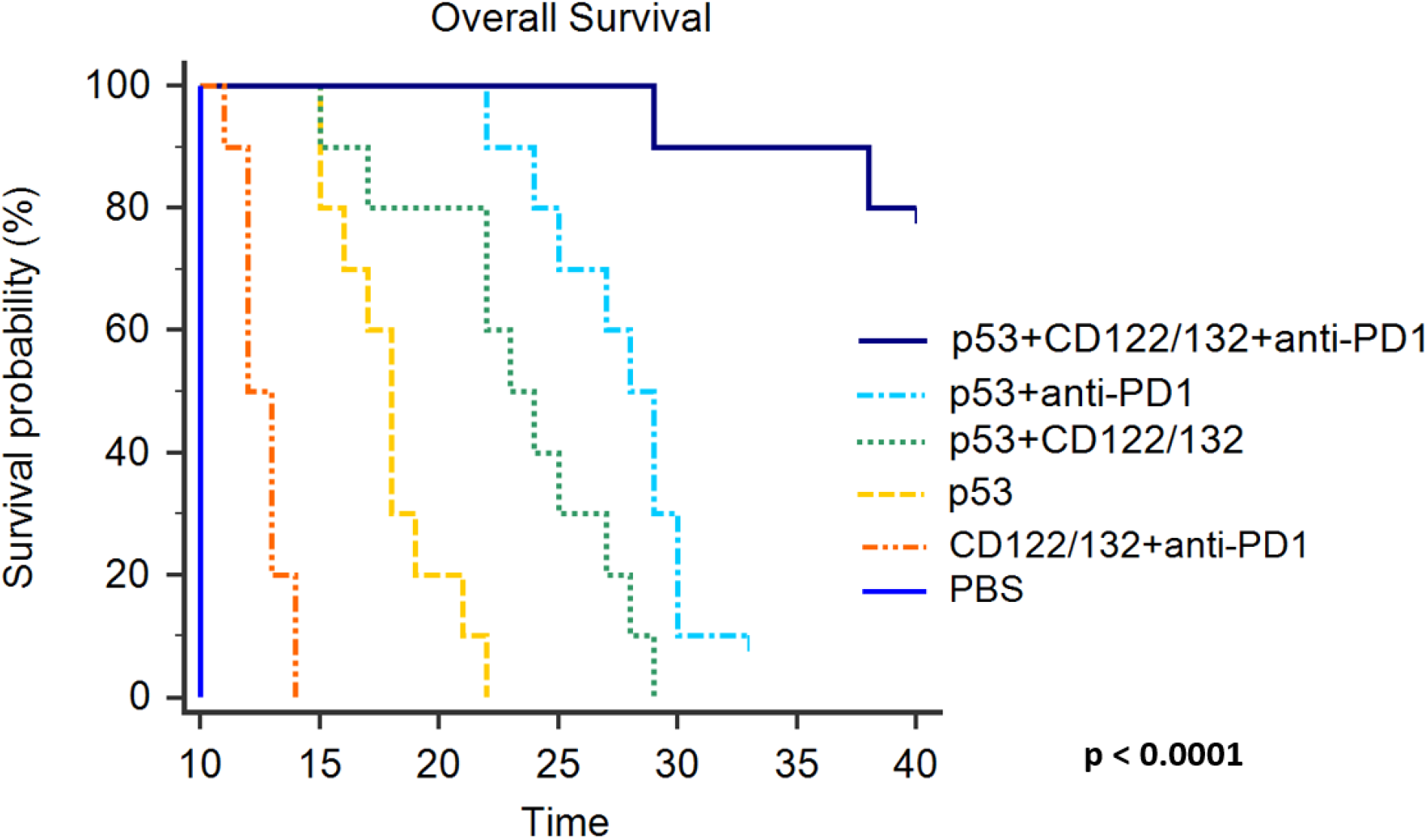
“Triplet” Ad-p53 + CD122/132 + anti-PD-1 treatment extends survival. Kaplan-Meier survival curves for mice treated with either PBS, CD122/132 + anti-PD-1, Ad-p53, or the combination of Ad-p53 + CD122/132, Ad-p53 + anti-PD-1 and Ad-p53 + CD122/132 + anti-PD-1. There was a statistically significant difference in these survival curves by the log rank test (p<0.0001 overall; p-value < 0.0003 comparing Ad-p53 + CD122/132 + anti-PD-1 treatment group vs. any other treatment group). These results also demonstrate an unexpected, substantial synergy of Ad-p53 + CD122/132 + anti-PD-1 therapy. The median survival of the Ad-p53 + CD122/132 + anti-PD-1 therapy group had not been reached after 40 days and 80% of this treatment group were still alive without evidence of any remaining tumors. In stark contrast, 98% (49/50) of animals in the other treatment groups had died by Day 30 and had median survivals ranging between 10 to 28 days.

In summary, these findings indicate that of all the Ad-p53 therapies, only the triplet combination Ad-p53 + CD122/132 + anti-PD-1 treatment resulted in potentially curative efficacy and long term survival by inducing synergistic local and systemic anti-tumor immunity with substantial abscopal effects.

### Gene expression profiles induced by Ad-p53 treatment

To assess the gene expression profiles most modulated by Ad-p53 treatment, mRNA isolated from pre and post Ad-p53 treatment biopsies were compared using the Nanostring IO 360 gene expression panel. The Nanostring IO 360 dataset was analyzed for genes substantially up- or down-regulated as a result of p53 treatment defined by a greater than or less than 10-fold change from baseline.

A total of 23 strongly-modulated genes out of the 770 gene set met these criteria. These genes with at least a 10 fold change in expression represented a highly, statistically significant gene subset most substantially effected by p53 treatment (p-value < 0.00001 by two-sided Fisher’s Exact Test compared to genes with less than a 10-fold change from baseline). These genes may be grouped into immune modulatory, stroma/fibrosis and tumor suppressor/cell cycle functional categories as listed in Table 1 and Figure 8. Unexpectedly, many of these genes were found to be involved in immune responses and anti-stroma/fibrosis functions which are not typically associated with p53 tumor suppressor mechanisms of action.

**Table 1.**
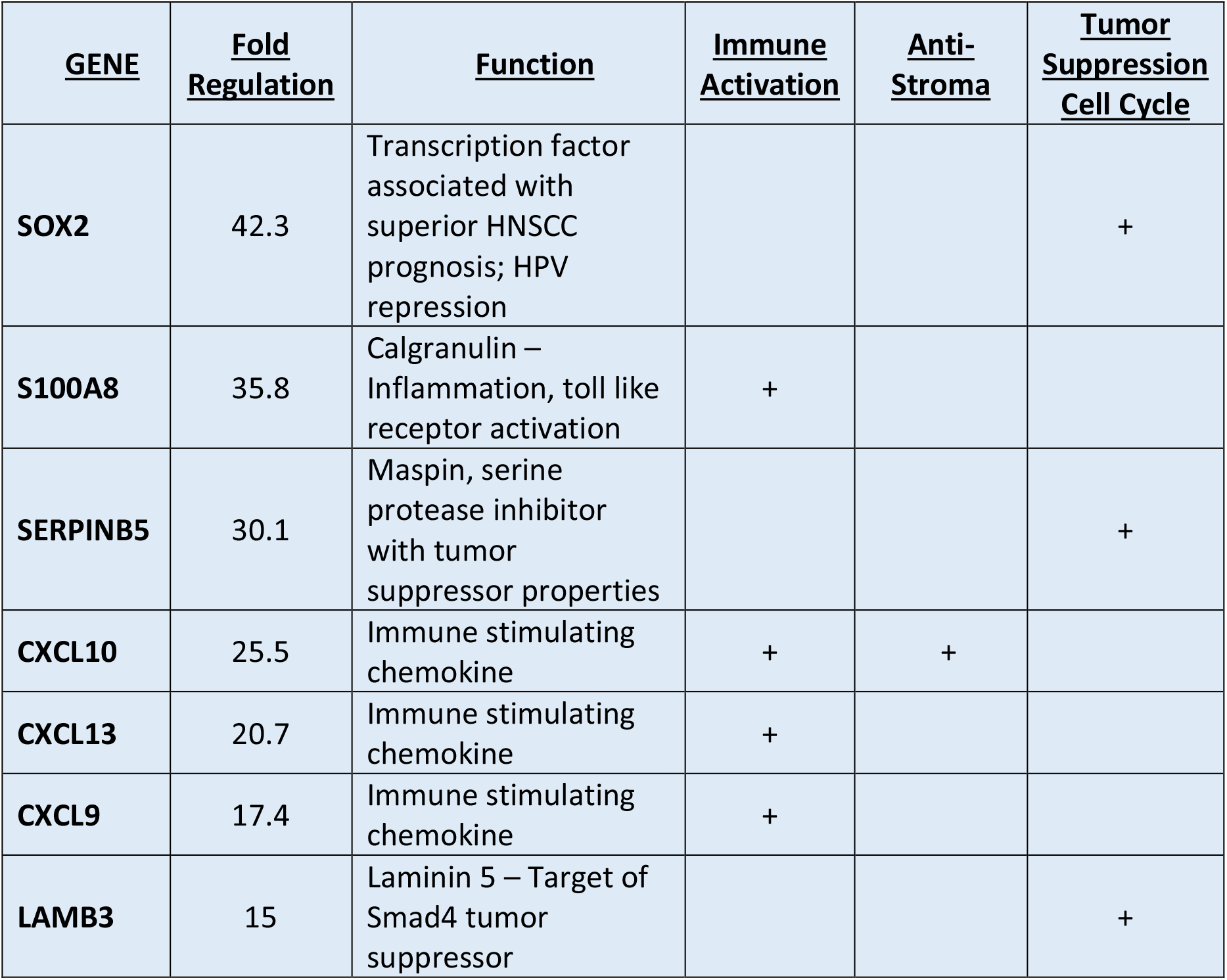
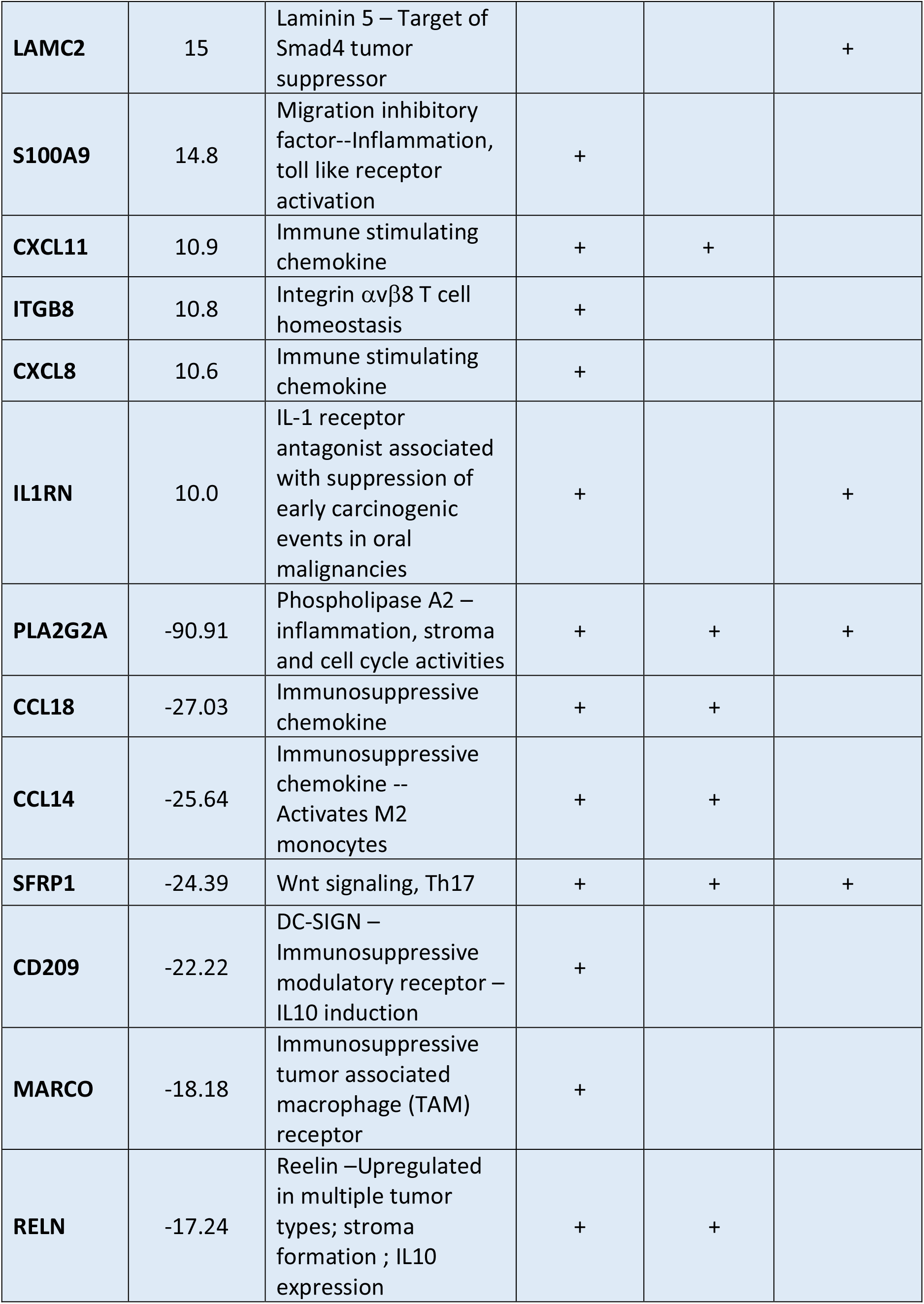
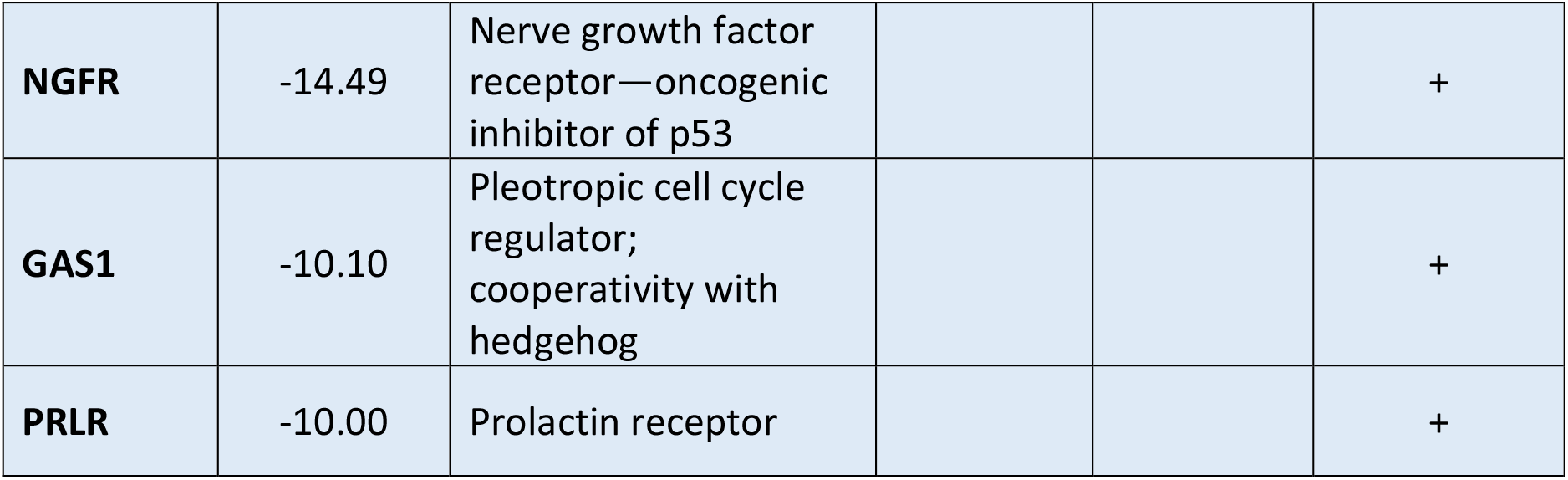

| **<u>GENE</u>** | **<u>Fold Regulation</u>** | **<u>Function</u>** | **<u>Immune Activation</u>** | **<u>Anti-Stroma</u>** | **<u>Tumor Suppression Cell Cycle</u>** |
| --- | --- | --- | --- | --- | --- |
| **SOX2** | 42.3 | Transcription factor associated with superior HNSCC prognosis; HPV repression |  |  | + |
| **S100A8** | 35.8 | Calgranulin – Inflammation, toll like receptor activation | + |  |  |
| **SERPINB5** | 30.1 | Maspin, serine protease inhibitor with tumor suppressor properties |  |  | + |
| **CXCL10** | 25.5 | Immune stimulating chemokine | + | + |  |
| **CXCL13** | 20.7 | Immune stimulating chemokine | + |  |  |
| **CXCL9** | 17.4 | Immune stimulating chemokine | + |  |  |
| **LAMB3** | 15 | Laminin 5 – Target of Smad4 tumor suppressor |  |  | + |
| **LAMC2** | 15 | Laminin 5 – Target of Smad4 tumor suppressor |  |  | + |
| **S100A9** | 14.8 | Migration inhibitory factor--Inflammation, toll like receptor activation | + |  |  |
| **CXCL11** | 10.9 | Immune stimulating chemokine | + | + |  |
| **ITGB8** | 10.8 | Integrin $\alpha v \beta 8$ T cell homeostasis | + | | |
| **CXCL8** | 10.6 | Immune stimulating chemokine | + |  |  |
| **IL1RN** | 10.0 | IL-1 receptor antagonist associated with suppression of early carcinogenic events in oral malignancies | + |  | + |
| **PLA2G2A** | -90.91 | Phospholipase A2 – inflammation, stroma and cell cycle activities | + | + | + |
| **CCL18** | -27.03 | Immunosuppressive chemokine | + | + |  |
| **CCL14** | -25.64 | Immunosuppressive chemokine -- Activates M2 monocytes | + | + |  |
| **SFRP1** | -24.39 | Wnt signaling, Th17 | + | + | + |
| **CD209** | -22.22 | DC-SIGN – Immunosuppressive modulatory receptor – IL10 induction | + |  |  |
| **MARCO** | -18.18 | Immunosuppressive tumor associated macrophage (TAM) receptor | + |  |  |
| **RELN** | -17.24 | Reelin –Upregulated in multiple tumor types; stroma formation ; IL10 expression | + | + |  |
| **NGFR** | -14.49 | Nerve growth factor receptor—oncogenic inhibitor of p53 |  |  | + |
| **GAS1** | -10.10 | Pleotropic cell cycle regulator; cooperativity with hedgehog |  |  | + |
| **PRLR** | -10.00 | Prolactin receptor |  |  | + |

**Figure 8.**
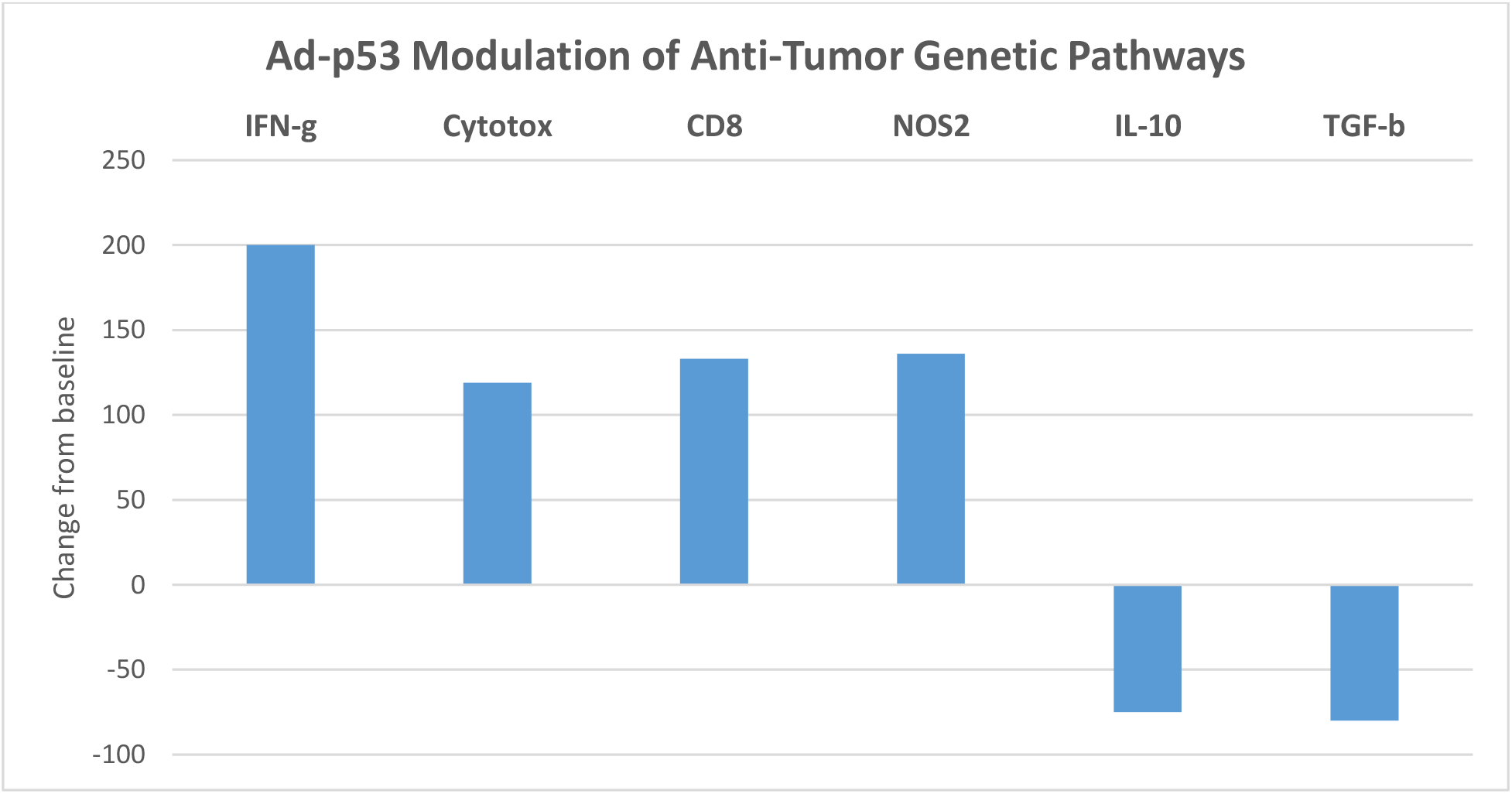
Concomitant Up Regulation of Immune Activating and Down Regulation of Immune Suppressive/Anti-Stromal Gene Pathways.

With respect to immune response modulating genes, expression of the proinflammatory S100A8 and S100A9 genes were up-regulated post-treatment by 35- and 15-fold, respectively. These genes are involved in pattern recognition receptor (PRR), damage associated molecular patterns (DAMPs), and pathogen-associated molecular patterns (PAMPs) which are key to the initiation of immune responses. The IFN gamma regulated chemokines CXCL8,9,10,11,13 were all up-regulated by 10->25-fold, reflecting their role in anti-tumor immune responses. The gene encoding Serpin B5 (maspin) was up-regulated by >30-fold and recent data indicate that maspin expression correlates with the activation and proliferation of CD8+ T-cell subsets and thus can modify the host immune response(16, 17).

In regard to the down regulation of gene expression contributing to increased anti-tumor immune responses, PLA2G2A which suppresses interferon-induced genes(18) had the greatest downregulation by >90-fold after Ad-p53 treatment. In addition, PLA2G2A is a direct target for beta-catenin-dependent Wnt signaling(19) and has been implicated in the regulation of Notch, TGF-beta and Hedgehog signaling pathways(19).The Wnt-beta-catenin and TGF-beta signaling pathways contribute to a lack of T cell infiltration in tumors and inhibits immune checkpoint blockade therapy(20, 21). The principal effector of the Wnt pathway, the CTNNB1 gene encoding beta-catenin was decreased by 3.6-fold, reflecting decreases in multiple components of beta-catenin signaling. Ad-p53 therapy resulted in a decrease in the immune suppressive chemokines CCL18 by >27-fold and CCL14 by >25-fold. CD209 (DC-SIGN), MARCO and RELN genes function in the down regulation of the immune system through IL10 and inhibitory tumor associated macrophage mechanisms respectively(22-24). CD209 is down-regulated by >20-fold, MARCO is down-regulated by >18-fold and RELN by > 17 fold.

Regarding anti-stromal/fibrosis effects, several chemokine genes associated with stoma/fibrosis formation were down regulated by 9 to 27 fold including CCL18, CXCL14, CXCL12. Another fibrosis-related gene is secreted frizzled receptor 1 (sFRP1) which was down-regulated by >24-fold. Multiple genes with anti-stromal/fibrosis effects were upregulated by Ad-p53 treatment. CXCL10 and CXCL11 are known to attenuate bleomycin induced pulmonary fibrosis CXCL10 and CXCL11 were increased by 25 and >10-fold, respectively, reflecting an anti-fibrotic activity of Ad-p53. Similarly, low levels of IL-1RN (IL-1 receptor antagonist) are associated with idiopathic pulmonary fibrosis and the IL-1RN gene was up-regulated by 10-fold.

The gene showing greatest up-regulation after Ad-p53 treatment was the transcription factor SOX2 (42-fold up regulation post-treatment). SOX2 (SRY-Box Transcription Factor 2) is associated with repressing tumorigenic HPV transcription(25). The gene encoding Serpin B5 (maspin) was up-regulated by >30-fold and has tumor suppressor anti-angiogenic functions. Other highly upregulated genes with tumor suppressor and/or cell cycle inhibitory activities are LAMB3, LAMC2, and IL1RN which were increased by 10 to 15 fold. PLA2G2A and SFRP1 which are associated with oncogenic cell cycling activity were down regulated by 90 and 24 fold respectively. Similarly, NGFR, GAS1 and PRLP have pleiotropic cell cycling properties and were inhibited by 14.49 to 10-fold following Ad-p53 treatment.

In addition to the individual genes in the Nanostring IO 360 dataset, the pre- and post-treatment biopsies data were also analyzed for gene signatures associated with immune activation, immune suppression and anti-stromal/fibrosis functions. As shown in Figure 8, Interferon-gamma, CD8+ T-cell profiles, Cytotoxicity and iNOS (inducible nitric oxide synthase, NOS2) profiles were increased consistent with activation of anti-tumor immune responses, whereas immunosuppressive pathways exemplified by IL-10 and TGF-beta and anti-stroma signatures were down regulated respectively.

Surprisingly, in addition to modulating immune mediators for anti-tumor immune responses, Ad-p53 therapy downregulated multiple gene pathways implicated in stroma/fibrosis formation. The stroma-related gene pathway (which comprises >50 gene products (see Supplemental Table 1) encompassing extracellular matrix remodeling, cell adhesion, myeloid cells, collagens, angiogenesis and metastasis was unexpectedly, strongly down-regulated by Ad-p53 treatment.

## Discussion

While immune checkpoint inhibitors are being increasingly employed in cancer treatment, most cancer patients do not respond to this form of immunotherapy(1). Similarly, genetically engineered versions of IL2 and IL15 cytokines with selective CD122/132 receptor activation have reduced toxicities and greater efficacy than their native proteins but the majority of tumors do not respond to these treatments either(2, 3). The syngeneic B16F10 melanoma is known to be resistant to these immunotherapies and is a useful model to explore novel immunotherapeutic approaches. In our studies, loco-regional Ad-p53 tumor suppressor gene therapy reversed resistance to both immune checkpoint inhibitor and selective CD122/CD132 IL2 and IL15 therapies, demonstrating unexpected synergies with abscopal effects on distant tumors that were not treated with Ad-p53. Remarkably, the “Triplet” therapy combining Ad-p53 with selective CD122/CD132 agonists and immune checkpoint blockade resulted in complete tumor remissions and potentially curative outcomes that significantly surpassed the efficacy of all other doublet and monotherapies tested which did not generate complete responses nor extended survivals.

These promising pre-clinical results led to the initiation of a Phase 1/2 clinical trial of combined Ad-p53 and anti-PD-1 therapy in patients with recurrent HNSCC. As reported elsewhere (Sobol R et al MedXriv), preliminary evaluation of pre and post Ad-p53 treatment biopsies were evaluated for changes in gene expression profiles and revealed increased Type I Interferon signaling, CD8+ T cell signaling and the Tumor Inflammation Signature which are all associated with increased responses to immune checkpoint inhibitors(14, 15, 26). Ad-p53 treatment also decreased known immune suppressive TGF-beta and beta-catenin signaling which may also contribute to enhanced immune checkpoint inhibitor efficacy.

In the present report, we examined the gene signatures associated with Ad-p53 treatment more thoroughly to provide additional insights into potential mechanism of actions for the observed synergies with IL2/IL15 agonists and immune checkpoint blockade. We identified 23 strongly-modulated genes with at least a 10 fold change in expression representing a highly, statistically significant gene subset most substantially effected by Ad-p53 treatment. These genes may be grouped into immune modulatory, stroma/fibrosis and tumor suppressor/cell cycle functional categories. Unexpectedly, many of these genes were found to be involved in immune responses and anti-stroma/fibrosis functions which are not typically associated with p53 tumor suppressor mechanisms of action.

With respect to immune response modulating genes, expression of the proinflammatory S100A8 and S100A9 genes were up-regulated post-treatment by 35- and 15-fold, respectively. These genes are involved in pattern recognition receptor (PRR), damage associated molecular patterns (DAMPs), and pathogen-associated molecular patterns (PAMPs) which are key to the initiation of immune responses(27). The IFN gamma regulated chemokines CXCL8,9,10,11,13 were all up-regulated by 10->25-fold, reflecting their role in anti-tumor immune responses. The gene encoding Serpin B5 (maspin) was up-regulated by >30-fold and data indicate that maspin expression correlates with the activation and proliferation of CD8+ T-cell subsets and thus can modify the host immune response(16, 17).

In regard to the down regulation of gene expression contributing to increased anti-tumor immune responses, PLA2G2A which suppresses interferon-induced genes(18) had the greatest downregulation by >90-fold after Ad-p53 treatment. In addition, PLA2G2A is a direct target for beta-catenin-dependent Wnt signaling(19) and has been implicated in the regulation of Notch, TGF-beta and Hedgehog signaling pathways(18). The Wnt-beta-catenin and TGF-beta signaling pathways contribute to a lack of T cell infiltration in tumors and inhibits immune checkpoint blockade therapy(20, 21). The principal effector of the Wnt pathway, the CTNNB1 gene encoding beta-catenin was decreased by 3.6-fold, reflecting decreases in multiple components of beta-catenin signaling. Ad-p53 therapy resulted in a decrease in the immune suppressive chemokines CCL18 by >27-fold and CCL14 by >25-fold. CD209 (DC-SIGN), MARCO and RELN genes function in the down regulation of the immune system through IL10 and inhibitory tumor associated macrophage mechanisms respectively(22-24). CD209 is down-regulated by >20-fold, MARCO is down-regulated by >18-fold and RELN by > 17 fold.

Surprisingly, in addition to modulating immune mediators for anti-tumor immune responses, Ad-p53 therapy downregulated multiple gene pathways implicated in stroma/fibrosis formation. The stroma-related gene pathway (which comprises >50 gene products (see Supplemental Table 1) encompassing extracellular matrix remodeling, cell adhesion, myeloid cells, collagens, angiogenesis and metastasis was unexpectedly, strongly down-regulated by Ad-p53 treatment. Several chemokine genes associated with stoma/fibrosis formation(28) were down regulated by 9 to 27 fold including CCL18, CXCL14, CXCL12. Another fibrosis-related gene is secreted frizzled receptor 1 (sFRP1)(29) which was down-regulated by >24-fold. Multiple genes with anti-stromal/fibrosis effects were upregulated by Ad-p53 treatment. CXCL10 and CXCL11 are known to attenuate bleomycin induced pulmonary fibrosis(30) and were increased by 25 and >10-fold respectively, reflecting anti-fibrotic activity of Ad-p53. Similarly, low levels of IL-1RN (IL-1 receptor antagonist) are associated with idiopathic pulmonary fibrosis(31) and the IL-1RN gene was up-regulated by 10-fold consistent with anti-fibrosis effects.

Regarding tumor suppressor/cell cycle inhibitory functions, the gene showing the greatest up-regulation after Ad-p53 treatment was the transcription factor SOX2 (42-fold up regulation post-treatment). SOX2 (SRY-Box Transcription Factor 2) is associated with repression of tumorigenic HPV transcription(25). The gene encoding Serpin B5 (maspin) was up-regulated by >30-fold and has tumor suppressor and anti-angiogenic functions(32). Other highly upregulated genes with tumor suppressor and/or cell cycle inhibitory activities are laminin-5 (LAMB3, LAMC2) (33) and IL1RN which were increased by 10 to 15 fold. PLA2G2A(18) and SFRP1(29) which are associated with oncogenic cell cycling activity were down regulated by 90 and 24 fold respectively. Similarly, NGFR(34), GAS1(35) and PRLP(36) have pleiotropic cell cycling properties and were inhibited by 14.49 to 10-fold following Ad-p53 treatment. Of particular relevance, NGFR is known to inhibit p53 and NGFR ablation enhances p53 activity(36).

In summary, loco-regional Ad-p53 tumor suppressor gene therapy reversed resistance to both immune checkpoint inhibitor and selective CD122/CD132 IL2 and IL15 therapies with substantial synergies. Remarkably, the “Triplet” therapy combining Ad-p53 with selective CD122/CD132 agonists and immune checkpoint blockade resulted in complete tumor remissions and potentially curative outcomes that significantly surpassed the efficacy of all other doublet and monotherapies tested none of which resulted in complete responses nor extended survivals. With respect to potential mechanisms of action, gene expression profiling comparing pre and post Ad-p53 tumor biopsies showed strong up-regulation of genetic pathways involved in anti-tumor immune responses including IFN-gamma activation, an increased CD8+ T cell signature, with concomitant down regulation of TGF-beta and IL10 gene profiles. Unexpectedly, Ad-p53 treatment substantially reduced fibrotic/stroma gene pathways. A number of previously unidentified, strongly p53 down regulated genes associated with stromal pathways and IL10 expression identified novel anti-cancer therapeutic applications. Ad-p53 treatment also decreased immune suppressive TGF-beta and beta-catenin signaling which may also contribute to enhanced immune checkpoint inhibitor efficacy. These mechanistic findings will need to be confirmed in a larger number of Ad-p53 treatment samples. Taken together, our initial results imply the ability of Ad-p53 to induce efficacious local and systemic anti-tumor immune responses with the potential to reverse resistance to immune checkpoint inhibitor therapy when combined with IL2 and IL15 CD122/132 agonists supporting the future clinical evaluation of this triplet therapy.

## Data Availability

The data supporting the conclusions of this article are either incorporated in the manuscript, its supplemental section or available by the corresponding author upon reasonable request.

## Acknowledgements

The authors wish to acknowledge the assistance and support of William B. Wells, Casey McCandless, Philip Seaton and Nicholas Puro. Without their efforts, this work would not have been completed.

## Author Contributors

RES, SC, KBM, and DW designed the research studies. HMA, B-KJ, C-OY and JN conducted the experiments. RES, SC, KBM, BS, MT and DW analyzed the data. RES, SC, KBM, and DW wrote the manuscript. All authors reviewed the results and approved the final version of the manuscript.

## Ethics Statement

The studies involving human participants were reviewed and approved by the ethics committee of the University of Toledo. The patients/participants provided their written informed consent to participate in this study.

All mice-related experiments were performed under an approved protocol following the institutional guidelines established by the institutional animal care and use committee (IACUC) of Hanyang University.

---

**Supplemental Table 1.**
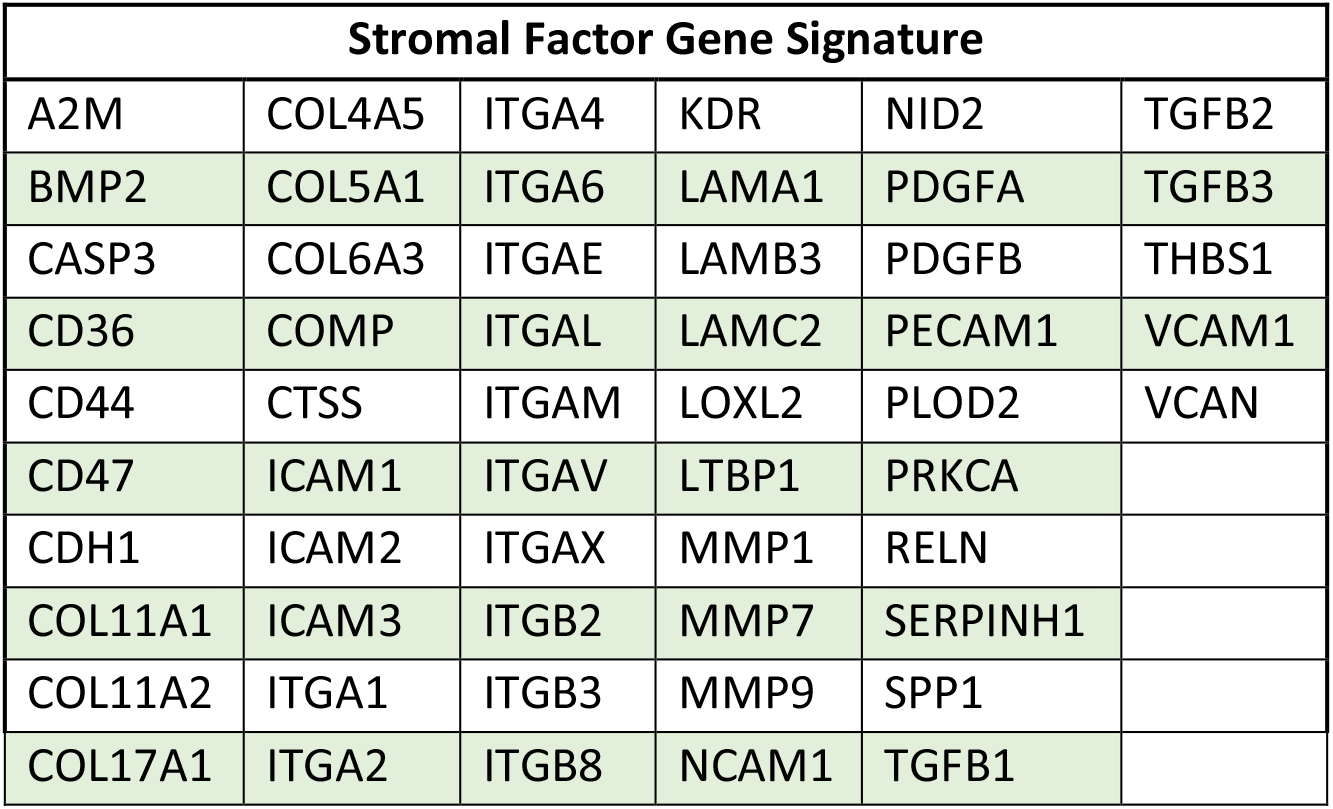
Stromal-related gene signature from IO360 panel.

**Supplemental Figure 1.**
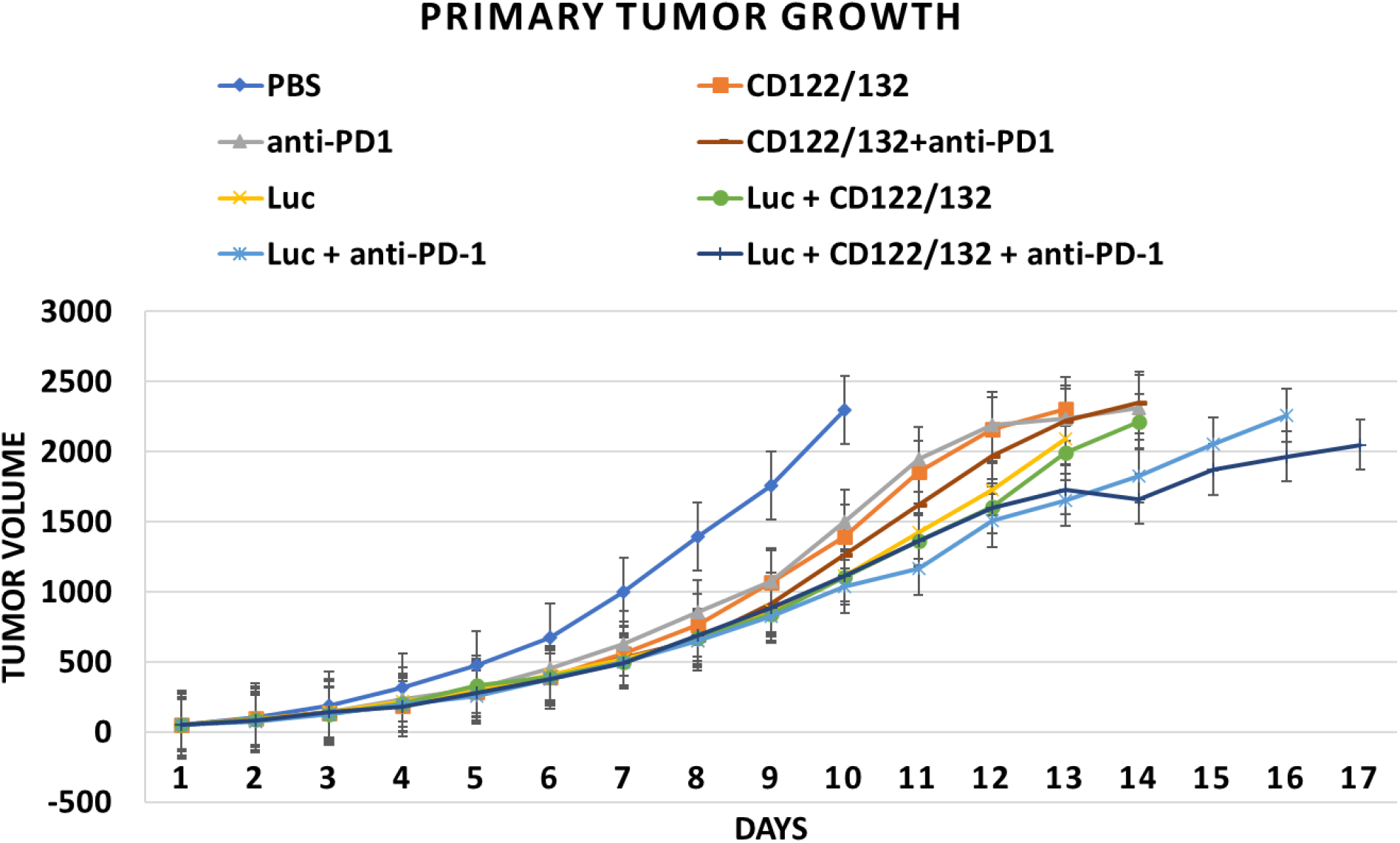
Ad-Luciferase (Ad-Luc) negative control + IL2 CD122/132 agonist + anti-PD-1 Efficacy: Tumor Volume. A graph showing primary tumor volumes over time in rodents receiving either phosphate buffered saline (PBS) control, CD122/132, anti-PD-1, IL2 CD122/132 + anti-PD-1, Ad-Luc control, or the combination of Ad-Luc control + IL2 CD122/132, Ad-Luc control + anti-PD-1 and Ad-Luc control + IL2 CD122/132 + anti-PD-1. In contrast to the treatments with Ad-p53, VirRx007 and Ad-IL24, there was no significant increase in therapeutic efficacy when Ad-Luc was combined with anti-PD-1, IL2 CD122/132, or IL2 CD122/132 + anti-PD-1 treatments. By day 16, the mean tumor volumes for all groups exceeded 2,000 mm^3^. A statistical analysis of variance (ANOVA) comparison of tumor volumes on Day 16 was not statistically significant (p-value = 0.1212; none of the mean tumor volumes between any of the treatment groups were statistically significant).

**Supplemental Figure 2.**
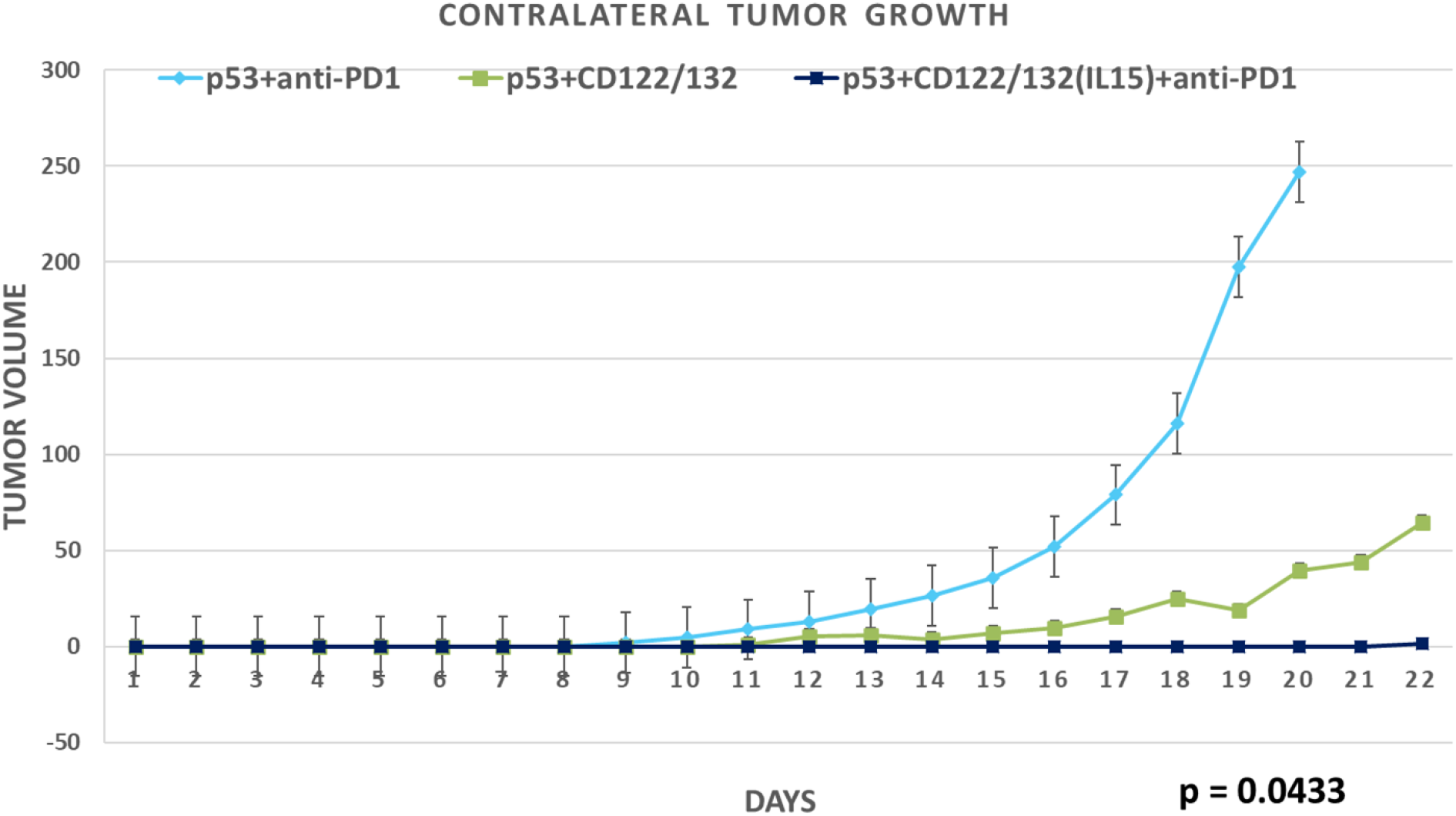
Substantially Superior Abscopal/Systemic Efficacy of “Triplet” Ad-p53 + IL15 CD122/132 agonist + anti-PD-1 Therapy. In this experiment, the preferential CD122/CD132 agonist was an immunocomplex comprised of recombinant IL15 and IL-15-R alpha-Fc. Consistent with the unexpected, substantially increased synergistic effects of Ad-p53 + IL15 CD122/132 + anti-PD-1 treatment on primary tumor growth, we also observed a surprisingly powerful and statistically significant abscopal effect of triplet Ad-p53 + IL15 CD122/132 + anti-PD-1 treatment compared to the other Ad-p53 treatment groups. A statistical analysis of variance (ANOVA) comparison of these contralateral tumor volumes determined synergy of the anti-tumor effects of Ad-p53 + IL15 CD122/132 + anti-PD-1 treatment (p-value = 0.0433 overall). Only the Ad-p53 + CD122/132 + anti-PD-1 group demonstrated a statistically significant decrease in contralateral tumor growth vs. the Ad-p53 + anti-PD-1 group (p-value = 0.0359).

