## Supplemental Table 1 for "Tumor Suppressor Immune Gene Therapy to Reverse Immunotherapy Resistance"

**Supplemental Table 1. Stromal-related gene signature from IO360 panel**

| Stromal Factor Gene Signature | | | | | |
|---|---|---|---|---|---|
| A2M | COL4A5 | ITGA4 | KDR | NID2 | TGFB2 |
| BMP2 | COL5A1 | ITGA6 | LAMA1 | PDGFA | TGFB3 |
| CASP3 | COL6A3 | ITGAE | LAMB3 | PDGFB | THBS1 |
| CD36 | COMP | ITGAL | LAMC2 | PECAM1 | VCAM1 |
| CD44 | CTSS | ITGAM | LOXL2 | PLOD2 | VCAN |
| CD47 | ICAM1 | ITGAV | LTBP1 | PRKCA | |
| CDH1 | ICAM2 | ITGAX | MMP1 | RELN | |
| COL11A1 | ICAM3 | ITGB2 | MMP7 | SERPINH1 | |
| COL11A2 | ITGA1 | ITGB3 | MMP9 | SPP1 | |
| COL17A1 | ITGA2 | ITGB8 | NCAM1 | TGFB1 | |
