## Supplemental Figure 1 for "Tumor Suppressor Immune Gene Therapy to Reverse Immunotherapy Resistance"

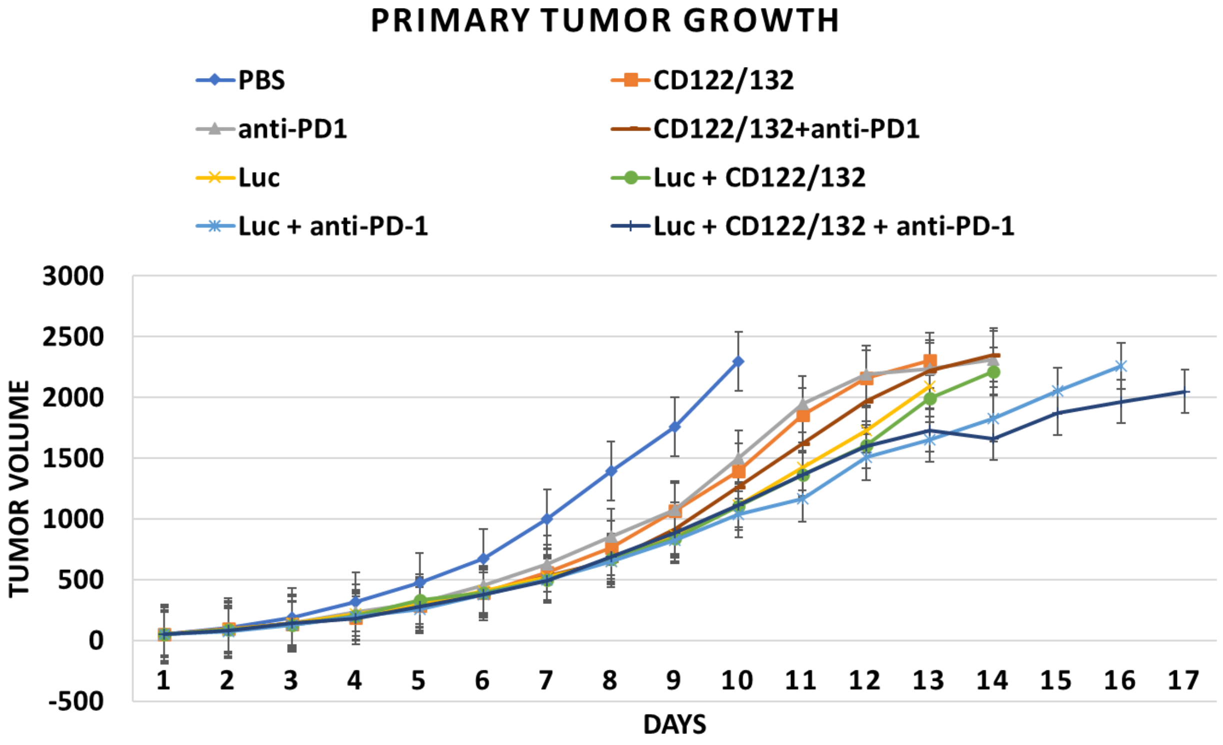


***Supplemental Figure 1. Ad-Luciferase (Ad-Luc) negative control + IL2 CD122/132 agonist + anti-PD-1 Efficacy: Tumor Volume.*** *A graph showing primary tumor volumes over time in rodents receiving either phosphate buffered saline (PBS) control, CD122/132, anti-PD-1, IL2 CD122/132 + anti-PD-1, Ad-Luc control, or the combination of Ad-Luc control + IL2 CD122/132, Ad-Luc control + anti-PD-1 and Ad-Luc control + IL2 CD122/132 + anti-PD-1. In contrast to the treatments with Ad-p53, VirRx007 and Ad-IL24, there was no significant increase in therapeutic efficacy when Ad-Luc was combined with anti-PD-1, IL2 CD122/132, or IL2 CD122/132 + anti-PD-1 treatments. By day 16, the mean tumor volumes for all groups exceeded 2,000* $mm^3$*. A statistical analysis of variance (ANOVA) comparison of tumor volumes on Day 16 was not statistically significant (p-value = 0.1212; none of the mean tumor volumes between any of the treatment groups were statistically significant).*
