## Supplemental Figure 2 for "Tumor Suppressor Immune Gene Therapy to Reverse Immunotherapy Resistance"

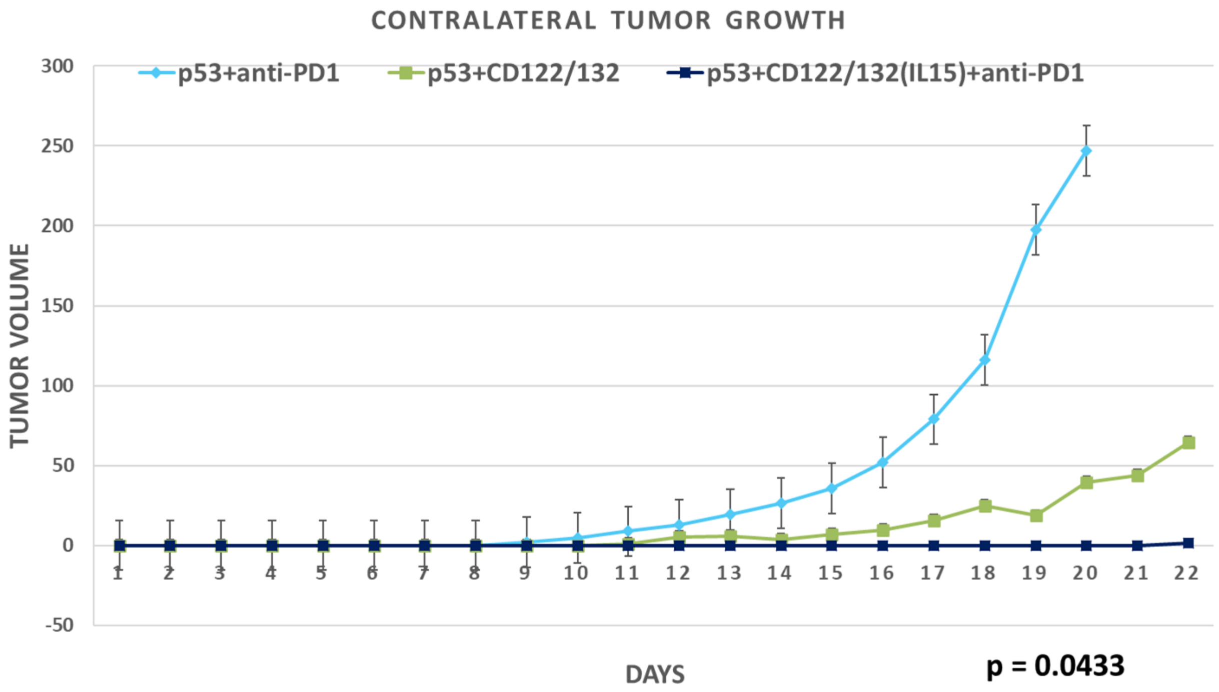


***Supplemental Figure 2. Substantially Superior Abscopal/Systemic Efficacy of "Triplet" Ad-p53 + IL15 CD122/132 agonist + anti-PD-1 Therapy.*** *In this experiment, the preferential CD122/CD132 agonist was an immunocomplex comprised of recombinant IL15 and IL-15-R alpha-Fc. Consistent with the unexpected, substantially increased synergistic effects of Ad-p53 + IL15 CD122/132 + anti-PD-1 treatment on primary tumor growth, we also observed a surprisingly powerful and statistically significant abscopal effect of triplet Ad-p53 + IL15 CD122/132 + anti-PD-1 treatment compared to the other Ad-p53 treatment groups. A statistical analysis of variance (ANOVA) comparison of these contralateral tumor volumes determined synergy of the anti-tumor effects of Ad-p53 + IL15 CD122/132 + anti-PD-1 treatment (p-value = 0.0433 overall). Only the Ad-p53 + CD122/132 + anti-PD-1 group demonstrated a statistically significant decrease in contralateral tumor growth vs. the Ad-p53 + anti-PD-1 group (*p-value = 0.0359*).*
